# A fully-automated integrative workflow to streamline NGS-based analyses within Molecular Tumour Boards

**DOI:** 10.64898/2025.12.12.25341897

**Authors:** Maria Giulia Carta, Miriam Angeloni, Lars Tögel, Christoph Schubart, Annett Hölsken, Robert Stoehr, Simona Vatrano, Davide Rizzi, Paolo Magni, Filippo Fraggetta, Arndt Hartmann, Florian Haller, Fulvia Ferrazzi

## Abstract

Molecular Tumour Boards (MTBs) rely on different bioinformatics tools and knowledgebases for variant annotation, oncogenicity classification, and estimation of complex biomarkers to identify actionable alterations. However, the typical bioinformatics workflow to process raw next-generation sequencing (NGS) data into clinically meaningful variants involves multiple steps and is inherently complex, thus requiring repeated manual intervention and causing delays in providing molecularly informed precision oncology. Here, we aimed at overcoming these limitations by developing a fully-automated integrative workflow to support NGS-based analyses within MTBs.

Our workflow was established at the Institute of Pathology, University Hospital Erlangen (Germany), and adapted to the fully digitized Pathology department at Gravina Hospital in Caltagirone (Italy), using the Illumina TruSight Oncology 500 HRD assay as case study. A trigger event initiates all the downstream bioinformatics analyses to support variant interpretation. In Erlangen, the trigger event is the automatic detection of new NGS data on the Illumina Connected Analytics cloud-based platform. In Caltagirone, the analyses are manually triggered from the anatomic pathology laboratory information system (AP-LIS).

The workflow automatically: (i) generates an intuitive overview of sequencing quality metrics, (ii) performs variant annotation, (iii) classifies variant oncogenicity through a fully-automated implementation of the ClinGen/CGC/VICC guidelines, and (iv) generates homologous recombination deficiency scores with genomic instability plots. In the digitized pathology department, results can be readily opened from the AP-LIS and visualized in the patient gallery.

Taken together, our end-to-end fully-automated workflow streamlines NGS-based analyses within MTBs by integrating variant interpretation, oncogenicity classification, and estimation of clinically relevant biomarkers.

## Introduction

In precision oncology, next-generation sequencing (NGS) plays a crucial role in identifying biomarkers that guide accurate diagnosis and inform treatment decisions [1]. Sequencing approaches, such as targeted gene panels, covering tens to hundreds of genes, whole exome sequencing, and whole genome sequencing are typically employed to capture tumour genomic features, which may provide molecularly informed personalised therapeutic strategies. While the technical and financial barriers to NGS data generation have been steadily diminishing [2], the translation of genomic data into actionable insights remains challenging, requiring extensive data analysis and expert interpretation. To align molecular findings with the available treatment options, Molecular Tumour Boards (MTBs) have been formed, consisting of a multidisciplinary team of oncologists, molecular pathologists, molecular biologists, and clinical bioinformaticians. The accurate and consistent interpretation of genomic data is central to the identification of clinically relevant alterations. This process can be summarised in three key steps: (i) variant functional annotation, i.e., the prediction of the potential effect of variants on gene transcripts and protein sequences [3], (ii) variant interpretation, i.e., the evaluation and summarization of the collected biomedical evidence supportive of this effect in the context of neoplastic disease [4], and ultimately (iii) variant classification as benign or oncogenic [5]. Each step typically requires running different bioinformatics tools, querying several different biomedical knowledgebases, and reconciling outputs across the various steps. Thus, manual intervention is unavoidable, leading to a fragmented and labor-intensive workflow. These technical barriers not only increase the workload of the working team, but might also prolong MTB preparation, potentially delaying treatment decisions [6]. In addition, owing to the lack of structured procedures as well as well-defined criteria for variant functional annotation and interpretation, institutions often adopt different strategies, which might lead to inconsistencies [7]. For example, variant interpretation can be approached either by using soft filters, whereby tags are added to the Variant Call Format (VCF) file without excluding any variants, or by applying hard filters that remove variants failing specific criteria. In addition, although internationally-recognised guidelines have been published to harmonise variant oncogenicity classification [8], there are very few publicly available tools that automate their assessment [9], thus further hindering consistent variant classification across clinical institutions. To promote the harmonisation of oncogenicity variant classification and encourage the adoption of the aforementioned guidelines for somatic variant classification [8], we have developed the Oncogenicity Variant Interpreter (OncoVI) (https://github.com/MGCarta/oncovi) [10], a tool that, based on their implementation, provides an automatic evaluation of variant oncogenicity.

In the present work, we aim to reduce molecular biologists’ workload and accelerate MTB preparation by proposing an end-to-end, fully-automated, integrative workflow to support NGS-based bioinformatics analyses within MTBs. Specifically, the developed workflow streamlines all the steps functional to clinical reporting, from the download of raw sequencing data to oncogenicity classification via OncoVI, and the estimation of clinically relevant biomarkers. Importantly, it only requires minimal, if any, user intervention. Using the TruSight Oncology 500 HRD assay as case study, the workflow was initially developed and established at the Institute of Pathology, University Hospital Erlangen (UKER) (Germany), and later adapted to the fully digitized pathology department at Gravina Hospital in Caltagirone (Italy), thus showing flexibility to different pathology environments with distinct informatics set-ups.

## Materials and Methods

### Tumour molecular profiling at the Institute of Pathology UKER before the fully-automated integrative workflow

At the Institute of Pathology UKER, patients can be enrolled in the MTB if they meet at least one of the following criteria: (i) advanced tumour disease with expected exhaustion of standard therapy within six months, (ii) cancer of unknown primary syndrome, or (iii) a tumour with an atypical course or the presence of multiple malignancies [11]. Starting from March 2025, all MTB patients have been systematically profiled using the TruSight Oncology 500 HRD (TSO500+HRD, Illumina, Inc., San Diego, CA, USA) gene panel covering 523 cancer-associated genes.

For tumour molecular profiling, the most recent formal-fixed paraffin-embedded (FFPE) tissue sample, if available, is used. More specifically, tumour tissue slices are cut from the FFPE block for histopathological evaluation and NGS analysis. After microdissection of the tumour tissue, DNA is isolated using standard techniques (Kit, Promega Maxwell) [11]. Library preparation follows the manufacturer’s instructions (https://support.illumina.com.cn/content/dam/illumina-marketing/apac/china/documents/tso500-hrd-data-sheet-m-gl-00748.PDF). Sequencing takes place in-house on the Illumina NextSeq500/550 sequencing platform and NGS data are processed using the DRAGEN TruSight Oncology 500 v2.6.0 Analysis Software deployed on the Illumina Connected Analytics (ICA) cloud platform. The platform employs a priority-based scheduling system, where analyses are queued and executed according to their priority levels [Low, Medium (default), or High] (https://help.ica.illumina.com/home/h-projects). On the ICA cloud platform FASTQ files are generated, reads are aligned to the human reference genome (hg19, UCSC University of California Santa Cruz), and variants (e.g., single- or multi-nucleotides variants, small insertions and deletions) are called.

Before the fully-automated integrative workflow was introduced, the NGS analyses outputs from the ICA cloud platform were downloaded by molecular biologists on a Linux-based server, where all the downstream bioinformatics analyses such as variant functional annotation and oncogenicity classification through OncoVI were manually launched. A pipeline developed for homologous recombination deficiency (HRD) score calculation, as well as a pipeline for the conversion of VCF files to Mutation Annotation Format (MAF) files, were also manually run at need.

### Design of the integrative workflow and implementation at the Institute of Pathology UKER

The fully-automated integrative solution was developed to streamline all the steps that, in the workflow outlined in the previous section, required manual intervention. It runs on a Linux-based server with Ubuntu 20.04.6 LTS operating system within dedicated conda environments (v24.11.1) to manage dependencies and ensure reproducibility.

A Python script (v3.10) runs continuously in the background and autonomously initiates the bioinformatics downstream analyses upon detection of a trigger event, thus ensuring a complete workflow automation. The script is executed in a detached screen session, remaining active in the background even after disconnection from the server. At the Institute of Pathology UKER, the trigger event is the presence of new NGS data on the ICA cloud platform, which is monitored by the Python script every 30 minutes. Upon detection of new NGS analyses outputs on the ICA cloud platform, the NGS data are automatically downloaded on the Linux-based server through *icav2* (v2.34.0), the command line interface for the ICA cloud platform (**Figure 1**).

**Figure 1.**
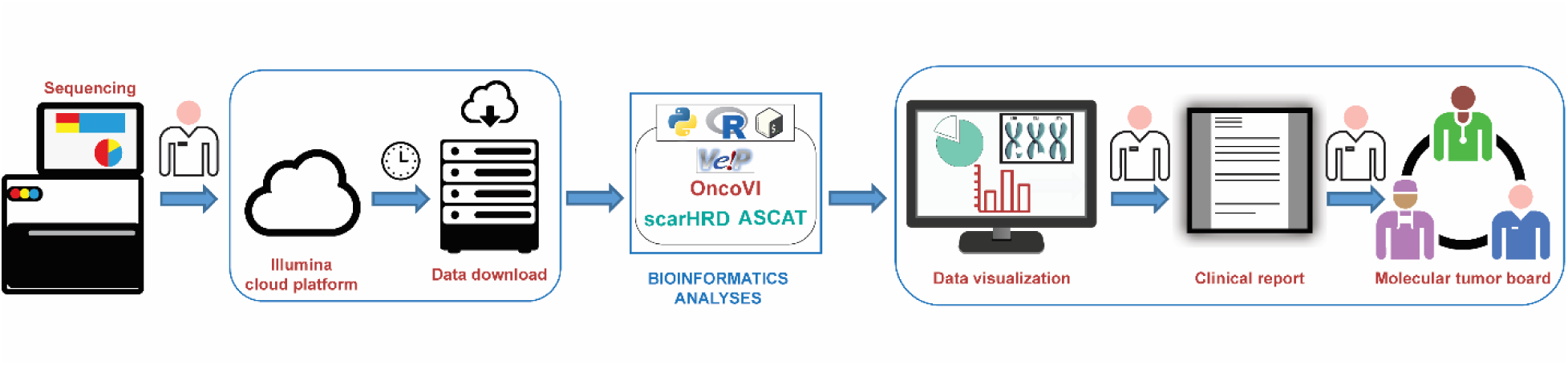
Configuration of the fully-automated integrative workflow at the Institute of Pathology UKER. At the Institute of Pathology UKER, a python script continuously monitors for new NGS data on the ICA cloud platform every 30 minutes. Upon detection of new NGS data, the script automatically downloads them via icav2, and executes all the downstream bioinformatics analyses, without manual intervention. Analysis outputs (plots, tabular data, and text files) are then used for clinical reporting by molecular biologists before discussion in the Molecular Tumour Board.

### Pipeline for variant interpretation

The integrative workflow includes: 1) a pipeline for variant interpretation, and 2) a custom pipeline for HRD score calculation. Prior to steps 1) and 2), quality control sequencing metrics are collected for all samples analysed within the same run (i.e., a batch of eight samples prepared and sequenced simultaneously with the TSO500+HRD assay). By default, the DRAGEN TSO500 Analysis Software generates, for each sample, a “MetricsOutput.tsv” file containing DNA library quality indicators (e.g., the percentage of bases with base quality >= 30, the percentage of aligned reads, the mean target coverage). To facilitate run-level evaluation, sample-level quality control sequencing metrics are aggregated into a comprehensive table and provided as Excel sheet.

The workflow proceeds with OncoVI for variant functional annotation, knowledge-based interrogation, and oncogenicity classification as previously described [10].

More specifically, first variants from the DRAGEN file “.hard-filtered.vcf” are functionally annotated relying on the Ensembl Variant Effect Predictor (VEP) (v113) [12] using as human reference genome the Ensembl build GRCh37. Then, they are enriched with existing biomedical information collected from several knowledgebases (e.g., ClinVar [13], COSMIC [14]). Ultimately, variants undergo oncogenicity classification through OncoVI [10], where each of them is assigned to one of five oncogenicity classes (i.e., “Oncogenic”, “Likely Oncogenic”, “Variant of Uncertain Significance (VUS)”, “Likely Benign”, “Benign”) along with a variant-specific score obtained as the sum of the points associated with the criteria triggered by OncoVI. A final table is produced in a human readable file format (.xlsx), containing all the variants enriched for the evidence collected during variant annotation and oncogenicity classification. This table provides molecular biologists with a structured and interpretable data set that serves as the basis for clinical interpretation and reporting.

Finally, the variants classified by the molecular biologists as Oncogenic, Likely Oncogenic, and VUS, together with the clinical data of the MTB patients, are integrated into a private instance of cBioPortal [15]. To format the patient associated variants for cBioPortal, the vcf2maf tool (v1.6.21) (Cyriac Kandoth. mskcc/vcf2maf: vcf2maf v1.6. (2020). doi:10.5281/zenodo.593251) is run. Specifically, the tool converts the annotated VCF file to a MAF file, a tab-delimited text file that associates each variant to its most clinically relevant gene transcript (typically the most severely affected). The resulting MAF file is then ready for integration into cBioPortal.

### Pipeline for HRD score calculation

The assessment of the HRD score was performed in R (v4.2.2) [16]. To this aim, we relied on (i) ASCAT (“Allele-Specific Copy Number Analysis of Tumors”) (v3.1.1) [17] for the inference of allele-specific copy number profiles of tumour only with a segmentation penalty set to 70, and (ii) scarHRD (v0.1.1) [18] for the calculation of HRD-associated genomic features [19, 20]. For each analysed sample, ASCAT uses the LogR ratios (LogR) and B-allele frequency (BAF) values generated by DRAGEN on the ICA cloud platform as input. The first represents a normalised measure of DNA copy number, while the latter is used to infer allelic imbalance. To predict sample germline genotypes, the ASCAT function *predictGermlineGenotypes()* is run providing customised parameters specific for the TSO500+HRD gene panel, not available in the original version of ASCAT. These parameters were derived through the evaluation of the variant allele frequency distribution of an exemplary case without copy number alterations, and set to the following values: proportionHetero= 0.29, proportionOpen= 0.03, and proportionHomo=0.68 so that their sum is 1. Additional parameters were set as follows: maxHomozygous= 0.05 and segmentLength= 100. The allele-specific copy number profiles generated by ASCAT are then used as input for scarHRD to estimate the HRD-associated genomic features, i.e., telomeric allelic imbalance (TAI) [21], loss of heterozygosity (LOH) [22], number of large-scale transitions (LST) [23], whose sum represents the total genomic instability score (GIS). The GIS, together with the three HRD-associated genomic features and the genomic instability plots produced by ASCAT, is provided in output by the integrative workflow.

### Adaptation of the integrative workflow to the pathology department at Gravina Hospital in Caltagirone

The developed integrative workflow was adapted to the Pathology department at Gravina Hospital in Caltagirone (Italy). Since 2019, the Italian pathology department has shifted towards a fully digitized diagnostic workflow and has implemented a fully tracked pathology system integrated within the pathology laboratory information system (AP-LIS) [24, 25].

Starting from November 2024, in Caltagirone tumour molecular profiling of cancer patients has been systematically performed either using the TruSight Oncology 500 (TSO500, Illumina, Inc., San Diego, CA, USA) or the TSO500+HRD gene panels. Analogously to the Institute of Pathology UKER, in the Italian pathology department DNA extraction, library preparation, and sequencing are executed in-house, with sequencing carried out on the Illumina NextSeq550 sequencing platform. However, differently from Erlangen, where NGS data processing is performed on the ICA cloud platform, in Caltagirone it takes place on a local Illumina DRAGEN server (v4.4.4) using the DRAGEN TruSight Oncology 500 Analysis Software. The output generated by the DRAGEN server is manually downloaded by molecular biologists on a network-attached storage mapped on a Linux-based server where afterwards all the downstream bioinformatics analyses take place (**Figure 2**).

**Figure 2.**
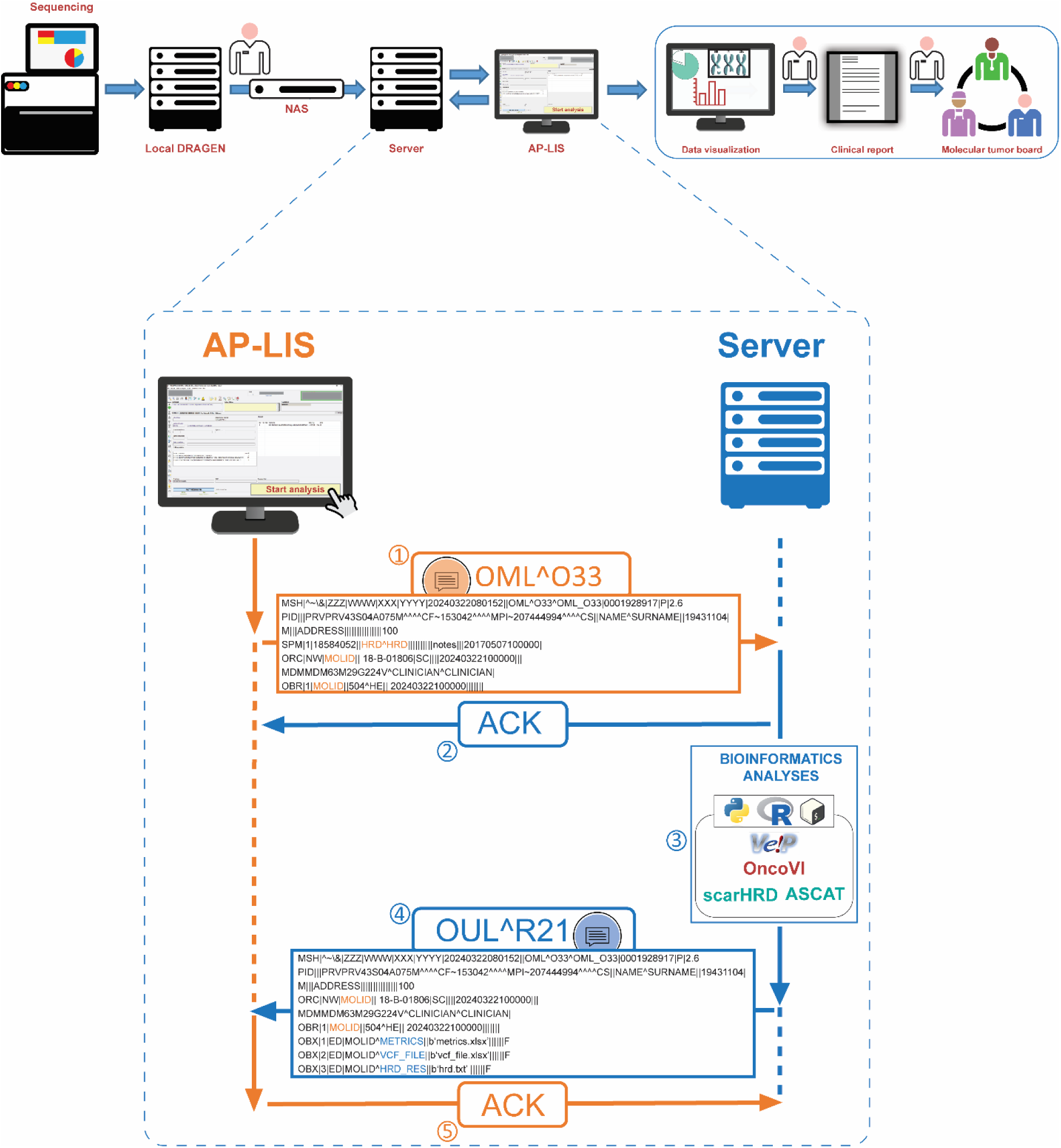
Adaptation of the fully-automated integrative workflow to the pathology department at Gravina Hospital. At the fully digitized pathology department at Gravina Hospital in Caltagirone, NGS data are manually retrieved from the Illumina DRAGEN server. Subsequent bioinformatics analyses for a given sample of a sequencing run can be requested by molecular biologists through the anatomic-pathology laboratory information system (AP-LIS) via Health Level 7 (HL7) messaging, which triggers the following sequence of events: (1) a laboratory order message (OML^O33) is sent from the AP-LIS to the remote server storing NGS data; (2) an acknowledgment (ACK) message is sent from the remote server to the AP-LIS upon reception of the OML^O33 message; (3) the remote server processes the OML^O33 message. First, the molecular identifier of the patient (orange) is retrieved, and all the downstream bioinformatics analyses are performed; (4) a laboratory observation message (OUL^R21) is sent from the remote server to the AP-LIS. Here, analysis results are transmitted to the AP-LIS as OBX segments (blue); (5) an ACK message is sent from the AP-LIS to the remote server upon reception of the OUL^R21 message. The figure panel depicting the HL7 connection between AP-LIS and server was adapted from [24].

The integrative workflow developed at the Institute of Pathology UKER, except for the vcf2maf conversion, was adapted to the fully digitized Italian pathology department. To this aim, a communication between the Linux-based server and the AP-LIS (Pathox v13.32.0, Tesi Elettronica e Sistemi Informativi S.P.A., Milan, Italy) was set-up via Health Level 7 (HL7) messaging through a Python-based server-client architecture as previously described [24]. Within this framework, the initiation of the bioinformatics analyses by the integrative workflow is triggered by an analysis request for a specific patient, submitted by molecular biologists through the AP-LIS (**Figure 2**).

This request is transmitted to the Linux-based server via an HL7 Laboratory Order Message (OML^O33). Once the analysis is completed, the results of the integrative workflow are transmitted from the Linux-based server to the AP-LIS via HL7 Unsolicited Laboratory Observation Message (OUL^R21) as OBX segments. Communication between the AP-LIS and the server is set-up via intranet connection using socket programming, and the HL7 messaging exchange takes place via a Transmission Control Protocol/Internet Protocol (TCP/IP) connection using the Minimal Lower Layer Protocol (MLLP) [24].

### Evaluation of hands-on time

The effectiveness in reducing the overall time required for downloading NGS data and performing downstream bioinformatics analyses of the fully-automated integrative workflow was evaluated at the Institute of Pathology UKER. To this aim, the following main steps of the standard molecular diagnostics workflow were identified:

1. downloading of NGS data from the ICA cloud platform to the Linux-based server used for downstream bioinformatics analyses;
2. execution of OncoVI and HRD score calculation pipelines;
3. generation of MAF files starting from VCF files.

Three molecular biologists working at the Institute of Pathology UKER (L.T., C.S., An.Hö.) recorded the hands-on time required for each step across five routine TSO500+HRD runs, each comprising data from eight patients. For comparison, the corresponding times required by the integrative workflow were considered. The Friedman test from the R package rstatix (v0.7.2) was utilized to assess differences in timing required by the three raters (molecular biologists) and required by the implemented fully-automated integrative workflow. Boxplots showing timing distribution across the three raters for each step were obtained relying on the R package ggplot2 (v3.5.2).

### User satisfaction survey design

To assess user satisfaction with the integrative workflow, a structured survey was conducted among the three biologists of the Institute of Pathology, UKER. The survey comprised a total of 12 statements aimed at evaluating molecular biologists’ perception of the workflow in terms of: (i) automation impact and results accessibility, (ii) efficiency, (iii) impact on work, and (iv) overall satisfaction (**Supplementary file 1**). Molecular biologists were asked to rate each statement using a Likert scale (ranging from “Strongly Disagree” to “Strongly Agree”), addressing factors such as reduced manual intervention, clarity of results availability, elimination of redundant steps, improved turnaround time for variant interpretation, decreased troubleshooting effort, and overall productivity gains. For each question and response category, the number of raters selecting that category (ranging from 0 to 3) was computed. The resulting 12 (questions) × five (Likert categories) matrix was visualized as a heatmap using the pheatmap R package (v1.0.13).

## Results

### Analysis outputs produced by the integrative workflow

The integrative workflow was developed to reduce manual workload and streamline MTBs preparation. At the Institute of Pathology UKER, patients are profiled with the TSO500+HRD assay in runs of eight tumour samples each, and analysed with DRAGEN TSO500 Analysis Software. A total of nine DRAGEN runs from the routine molecular diagnostics were analysed through the integrative workflow from July, 19^th^ 2025 to October, 22^nd^ 2025. The average time taken by the workflow for the analysis of a sequencing run (from download of NGS analyses outputs from the ICA cloud platform to the final generation of plots and tabular data) was 26.8 minutes.

To provide molecular biologists with an overview of the sequencing metrics quality control, the metrics of all samples belonging to the same sequencing run are collected into a unique tabular file, called “Metrics_Summary.xlsx” (**Supplementary Figure 1**). In this table, rows correspond to the metrics calculated by DRAGEN and columns represent the analysed samples. A cell will be assigned a “PASS” flag if the metric it contains falls within the threshold set by DRAGEN, otherwise the cell will be assigned a “FAILED” flag and automatically marked in red.

After quality control, OncoVI, a tool for variant oncogenicity classification [10], and the pipeline for HRD score calculation are executed. The bioinformatics workflow for variant functional annotation and knowledgebases interrogation included in OncoVI produces an Excel annotation file with the extension “_prediction_vep.xlsx”. In this file, each row corresponds to a variant that passed all the filters applied in the variant annotation step, while the columns provide curated information from the interrogated knowledgebases. A separate tabular file with extension “_OncoVI_prediction.xlsx” is also produced to store variants oncogenicity evaluation performed by OncoVI. This file reports for each variant included in the “_prediction_vep.xlsx” file the variant-specific score, the oncogenicity classification, and the criteria triggered by OncoVI that contribute to the variant-specific score. Once OncoVI assessment is completed, the generation of the MAF files takes place using the “filter_vep_samplename_dna_filtered.vcf” file as input, which is the VCF file annotated by Ensembl VEP before the conversion to the human readable format.

After the generation of the MAF files, the pipeline for HRD score calculation is executed and the plots generated by ASCAT are made available to molecular biologists to support the evaluation of patient tumour genomic instability. These plots include: (i) the LogR and BAF tracks, (ii) the allele-specific copy number profile of the tumour, and (iii) the sunrise plot (**Supplementary Figure 2**). Specifically, the sunrise plot shows tumour purity and ploidy estimates generated by ASCAT. The green cross corresponds to the best purity-ploidy combination identified by ASCAT that fits the observed LogR and BAF data. Furthermore, the total HRD score (GIS) as well as TAI, LOH, and LST values are made available in the text file named “HRDresults.txt”.

The framework was also integrated in the fully digitized Italian pathology department at Gravina Hospital in Caltagirone relying on HL7 messaging. Here, a total of five DRAGEN runs, of eight patients each, from the routine molecular diagnostics were analysed through the integrative workflow between July, 21^st^ 2025 and October, 20^th^ 2025. The average time taken by the workflow for the analysis of a sequencing run (from analysis request from the AP-LIS to the final generation of plots and tabular data) was 12.4 minutes. In Caltagirone, analysis results are directly accessible by molecular biologists from the AP-LIS (**Figure 3**).

**Figure 3.**
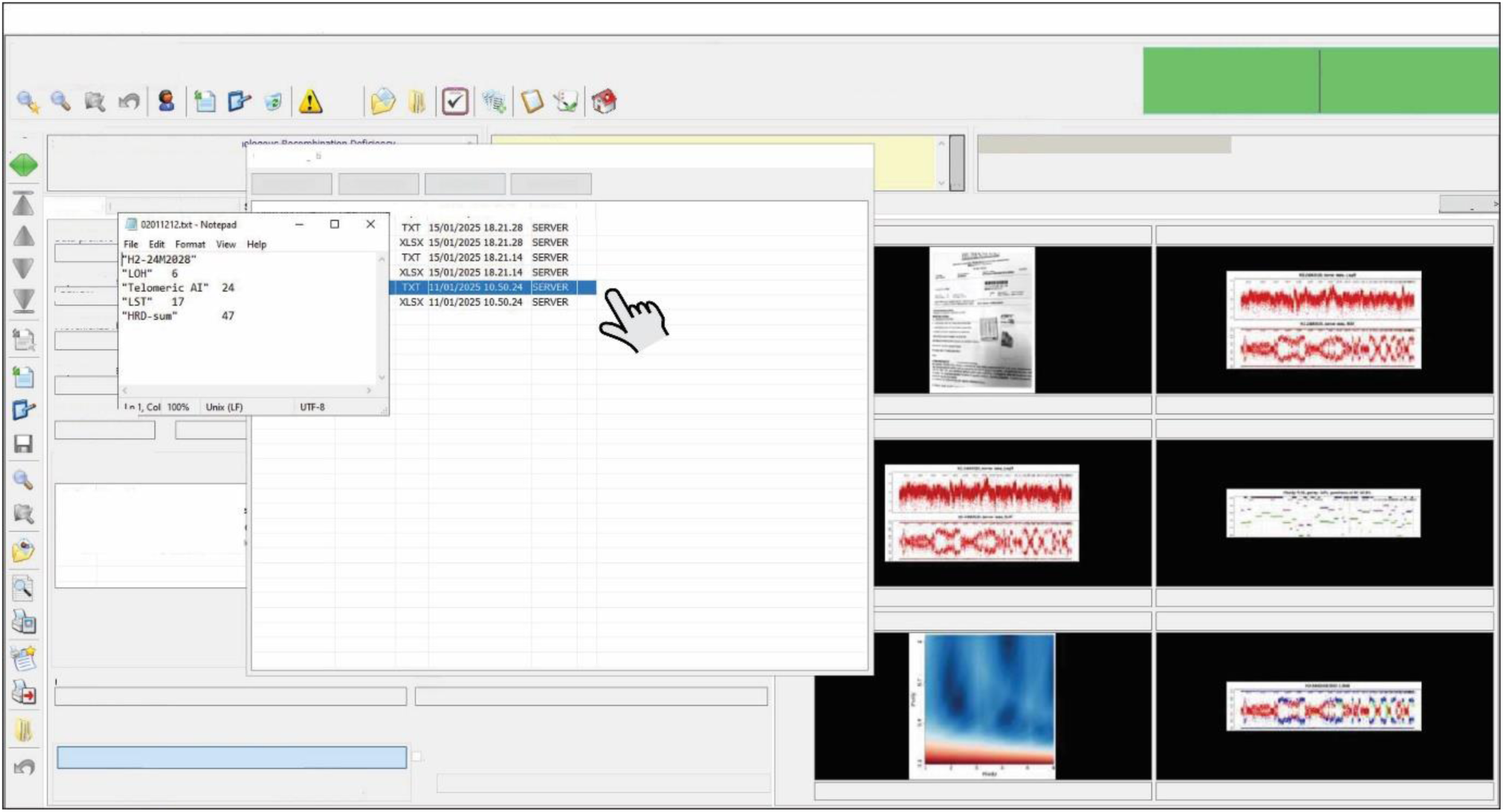
Visualisation of the output produced by the integrative workflow in the AP-LIS. Example of analysis outputs visualisation in the anatomic pathology laboratory information system (AP-LIS) of the pathology department at Gravina Hospital in Caltagirone. The text and tabular files generated by the workflow are available as attachments and can be opened with a double click (left), whereas all the plots are made available in the patient’s gallery (right).

Taken together, the developed integrative framework allows a fully automated execution of all the typical downstream bioinformatics analyses, usually manually performed by molecular biologists in the context of MTBs. In addition, it provides flexibility for integration in pathology environments with different informatics set-ups.

### Impact of the automated workflow on bioinformatics analyses execution time

To systematically evaluate the effectiveness of the workflow, the time taken by the three molecular biologists of the Institute of Pathology UKER to: (1) download NGS data from the ICA cloud platform to the Linux-based server, (2) run OncoVI and HRD score calculation pipelines, and (3) generate MAF files was compared with the execution time of the automated workflow. This assessment was conducted for five independent TSO500+HRD routine sequencing runs.

The molecular biologists took a median time of 12.73, 10.33, and 14.55 minutes, respectively, across the five runs for the manual download of raw NGS analyses outputs from the ICA cloud platform to the local server used for downstream bioinformatics analyses (**Figure 4a**). In contrast, the integrative workflow required a median time of 9.69 minutes. This difference was statistically significant (Friedman test, p = 0.026), thus highlighting the efficiency of the automated approach in the data acquisition phase.

**Figure 4.**
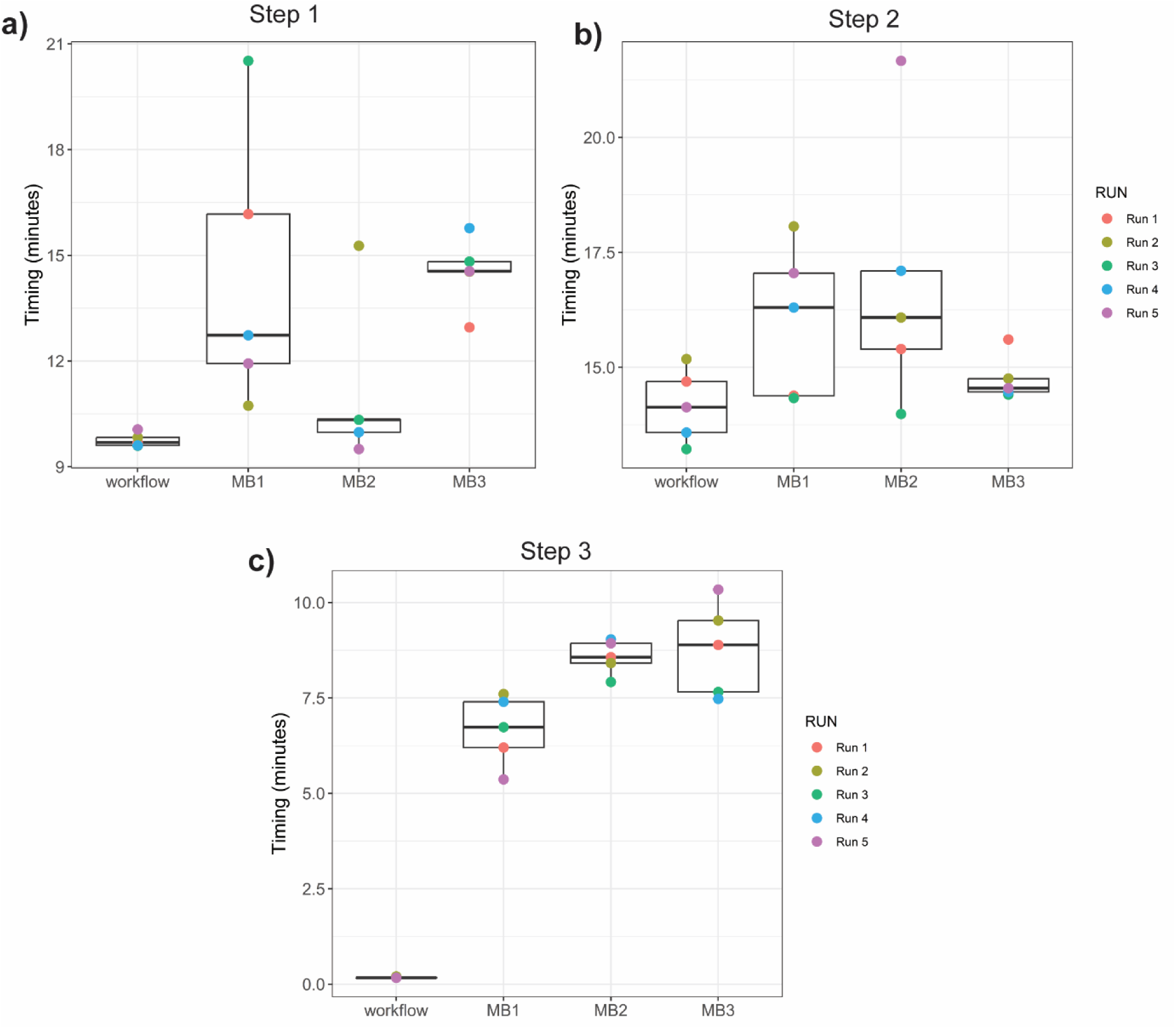
Comparison of execution times between the automated workflow and the three molecular biologists at the Institute of Pathology UKER. Boxplot distribution of the time registered across five different sequencing runs to: **a)** download NGS analyses outputs from the ICA cloud platform to the Linux-based server, **b)** run the downstream bioinformatics analyses on the Linux-based server, and **c)** generate MAF files starting from VCF file. MB = molecular biologist.

To run OncoVI and the HRD score calculation pipeline, the execution time of the integrative workflow was in median 14.13 minutes across the five sequencing runs, while the molecular biologists required a median time of 16.30, 16.08, and 14.55 minutes, respectively. Although the automated framework demonstrated a slight advantage, the difference was not statistically significant (Friedman test, p = 0.14) (**Figure 4b**).

The third step, involving the vcf2maf conversion, showed a marked difference between manual and automated execution. Indeed, the biologists spent a median time of 6.73, 8.57, and 8.89 minutes, respectively, across the five runs. In contrast, the integrative workflow took a median time of 0.17 minutes. The Friedman test confirmed the statistical significance (Friedman test, p = 0.0036), underscoring the substantial gain provided by the automated approach (**Figure 4c**).

Collectively, these results show how the integrative workflow conferred tangible improvements in terms of time efficiency compared to the manual analysis steps.

### Survey results

The three molecular biologists of the Institute of Pathology UKER, were asked to participate in a survey aimed at assessing their satisfaction with the routine adoption of the fully-automated integrative workflow (**Supplementary file 1**). When evaluating the impact of workflow automation and the accessibility of the results (**Figure 5**), all participants agreed that the automated framework requires no human intervention and delivers results in an intuitive way. With respect to workflow efficiency, the participants strongly agreed that the automated workflow reduces repetitive tasks required in the non-automated solution and saves time during variant interpretation. Regarding the impact on their daily work, a positive effect on their overall productivity was perceived. Indeed, participants reported that neither troubleshooting nor continuous monitoring of the analysis was longer needed. Finally, in terms of overall satisfaction, all respondents indicated that they were highly satisfied with the benefits gained with the integrative workflow, and experienced a significant improvement. Moreover, all participants recognized the great benefit for other colleagues in the field.

**Figure 5.**
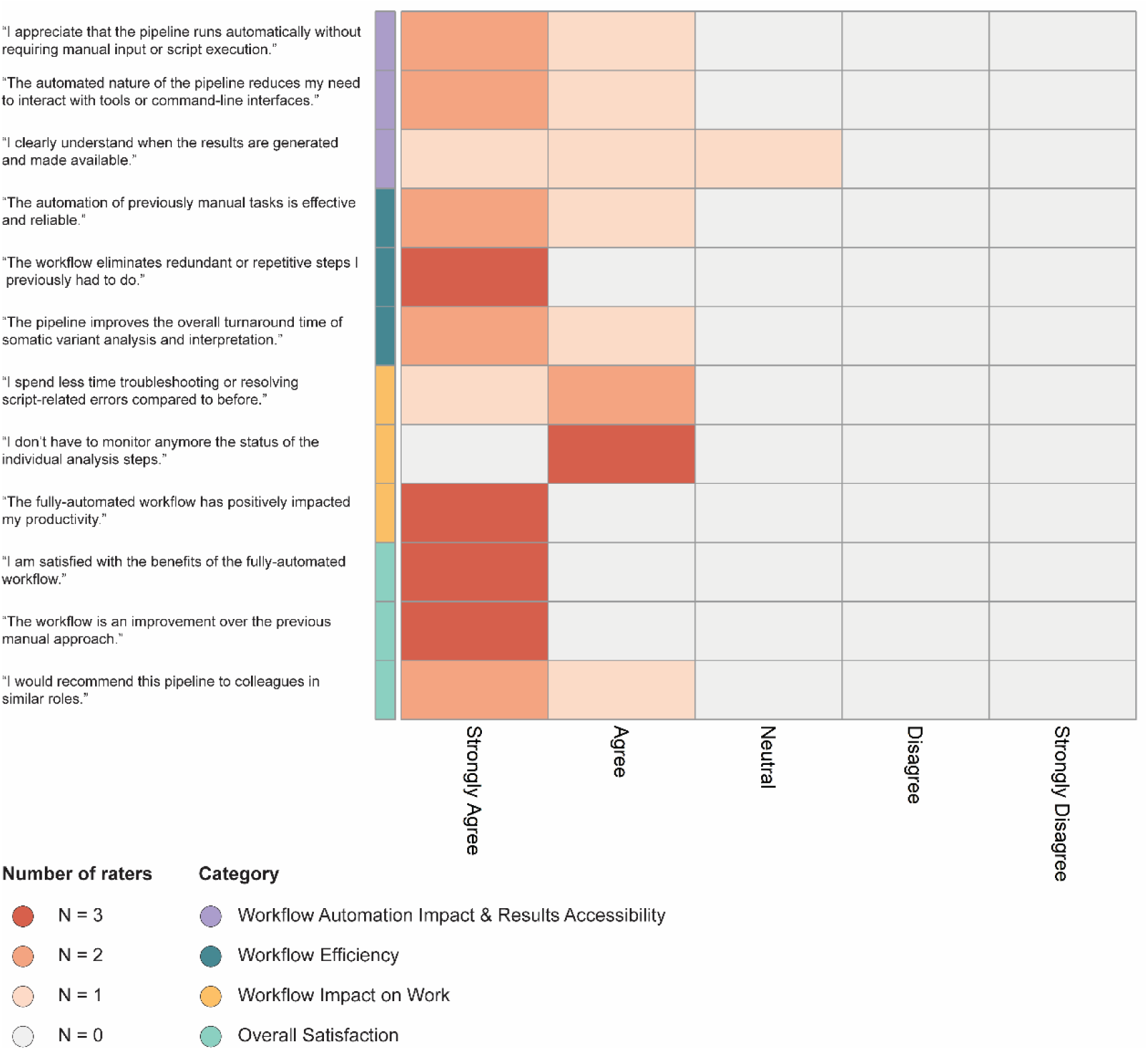
Heatmap representation of survey responses. For each question (rows) and response category (Strongly Agree-Strongly Disagree) (columns), the colour intensity reflects the number of raters (0–3) who selected a given Likert response for each question, with light grey indicating 0 raters and dark red indicating 3 raters.

Collectively, these findings demonstrate that the implementation of the integrative workflow in the clinical practice was well-accepted by the users, enhanced overall efficiency, and showed a positive impact on their workload.

## Discussion

Difficulties in achieving reproducible and standardised interpretation of somatic variants from NGS-based tumour molecular profiling have been widely recognized [26]. Notably, tumour molecular profiling provides the technical foundation for precision oncology, yet downstream analyses and interpretation of NGS data remain challenging. Expert manual curation is central to high-quality clinical interpretation. However, the process is often slowed down by repetitive, manual tasks. In addition, the scarcity of automated solutions based on standardized guidelines makes the whole process highly variable across institutions. Thus, there is a pressing need to develop automated and standardised solutions to improve reproducibility, reduce manual workload, and accelerate MTBs preparation.

Here, we presented the implementation and evaluation of an end-to-end, fully-automated integrative workflow that streamlines and harmonises NGS-based analyses within MTBs. Using the TSO500+HRD gene panel as case study, our integrative solution was initially designed to streamline the variant interpretation workflow at the Institute of Pathology UKER, and later adapted to a fully digitized Italian pathology department at Gravina Hospital in Caltagirone. The developed workflow allows minimizing, if not completely removing, the need for manual intervention throughout the execution of typical downstream bioinformatics analyses for variant interpretation.

The outputs provided by our integrative strategy include: (i) an intuitive overview of sequencing metrics across patients within a run, (ii) standardised oncogenicity classification of somatic variants via OncoVI, and (iii) the generation of plots and tabular data supporting data curation and interpretation by molecular biologists.

When we evaluated the time savings achieved by the integrative workflow, we observed a substantial reduction of the time required for data analysis compared to the manual approach. A recent study on digital strategies supporting MTBs workflows has shown how automation reduces preparation and discussion times, standardises documentation, and facilitates case presentation [6]. In line with these findings, our workflow demonstrates how automated solutions can free experts from repetitive and error-prone tasks (e.g., reformatting, copying and pasting commands for tool execution), thus enabling them to focus on higher-level activities like data curation and variant interpretation. This was confirmed by the survey conducted at the Institute of Pathology UKER, where the workflow’s user-friendliness, efficiency, and impact in significantly reducing the workload were identified by responders as the main benefits.

We also showed that our integrative solution can be adapted to working environments with different infrastructures. Indeed, its deployment in both analogue and fully digitized pathology departments further highlighted its versatility in real-world settings. In Caltagirone’s pathology department, the output of our integrative strategy could be directly visualized within the AP-LIS, thus ensuring a convenient and rapid access to well-organised results for quality control, variant interpretation, and HRD score calculation. This emphasizes the benefits of having a seamless interoperability with the AP-LIS. Enhancing and promoting interoperability would be desirable also in other pathology departments. Indeed, this would first and foremost allow a more convenient access to results directly from patient records. Secondly, it would facilitate the connection of workflow output with additional digital solutions useful in MTB settings. Tools such as cBioPortal [15, 27, 28] already facilitate genomic data integration and visualisation for treatment recommendations in patients with similar mutational profiles [29]. Yet, they require structured and standardised input. A fully automated strategy could help in gaining efficiency and scalability, as well as facilitating the adoption of harmonised data formats to fully exploit the benefits of such tools [6]. In an era where the volume of NGS data is constantly growing and where data are combined with associated clinical information, the development of digital solutions and visualisation strategies that can support data integration, reuse, and efficient interpretation is of paramount importance [26, 30, 31].

It is worth mentioning that the evaluation of our strategy was limited to a small number of sequencing runs. In addition, runtime analysis across varying hardware configurations as well as different physical computing resources was not explored.

In conclusion, the presented integrative solution significantly reduces molecular biologists’ manual efforts, accelerates turnaround times, and shifts the focus of experts towards variant interpretation and clinical reporting. Importantly, it represents a concrete step towards reducing subjectivity, improving reproducibility, and embedding automated workflows within routine clinical frameworks. In addition, by generating results in harmonised data formats (e.g., MAF file), our integrative strategy can favour interoperability with platforms such as cBioportal, eliminating the need for manual transformations to align analysis’ outputs with platform prerequisites. Taken together, we believe that our workflow offers a valuable support to streamline NGS-based analyses within MTBs, with the potential to be extended to more diverse type of NGS data analyses and hospital settings.

## Author Contributions

M.G.C., M.A., and Fu.Fe. conceived the study, interpreted the data, and wrote the manuscript; M.G.C. implemented the pipelines for variant interpretation and HRD score calculation; M.A. implemented the workflow in the pathology department at Gravina Hospital in Calagirone; L.T., C.S., and An.Hö. tested the workflow for the hands-on-time evaluation and participated in the survey; D.R. embedded results in Pathox; R.S., S.V., P.M., Fi.Fr., Ar.Ha., and F.H. contributed materials, medical, and/or methodological expertise. All authors have read and approved the final manuscript. We are grateful to the teams of the Molecular Diagnostics Laboratory of the Institute of Pathology UKER, of the Zentrum für Personalisierte Medizin Erlangen, and of the Molecular Tumour Board Erlangen.

## Data availability statement

All data produced in the present work are contained in the manuscript.

## Ethics approval and consent to participate

The study was conducted in accordance with the Declaration of Helsinki and approved by the Ethics Committee of the Friedrich-Alexander University Erlangen-Nuremberg (100_17 B from 7 April 2017, addendum from 27 July 2021).

---

**Supplementary Figure 1.**
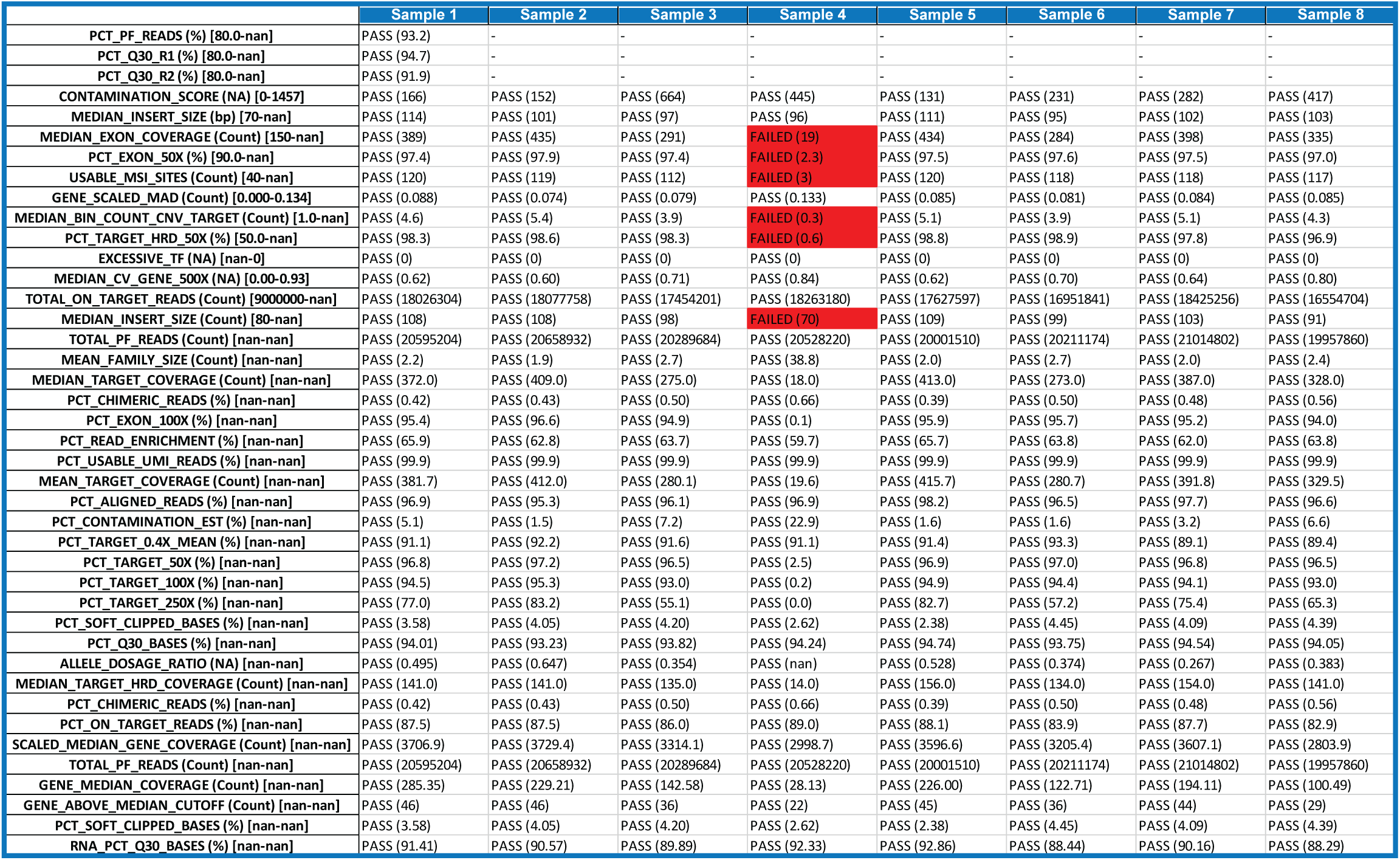
Overview of the metrics summary file. Example of a metrics summary file (“Metrics_Summary.xlsx”) storing the sequencing metrics, as calculated by DRAGEN, for all the samples belonging to the same sequencing run. Metrics that failed quality thresholds according to the imposed by DRAGEN are highlighted in red.

**Supplementary Figure 2.**
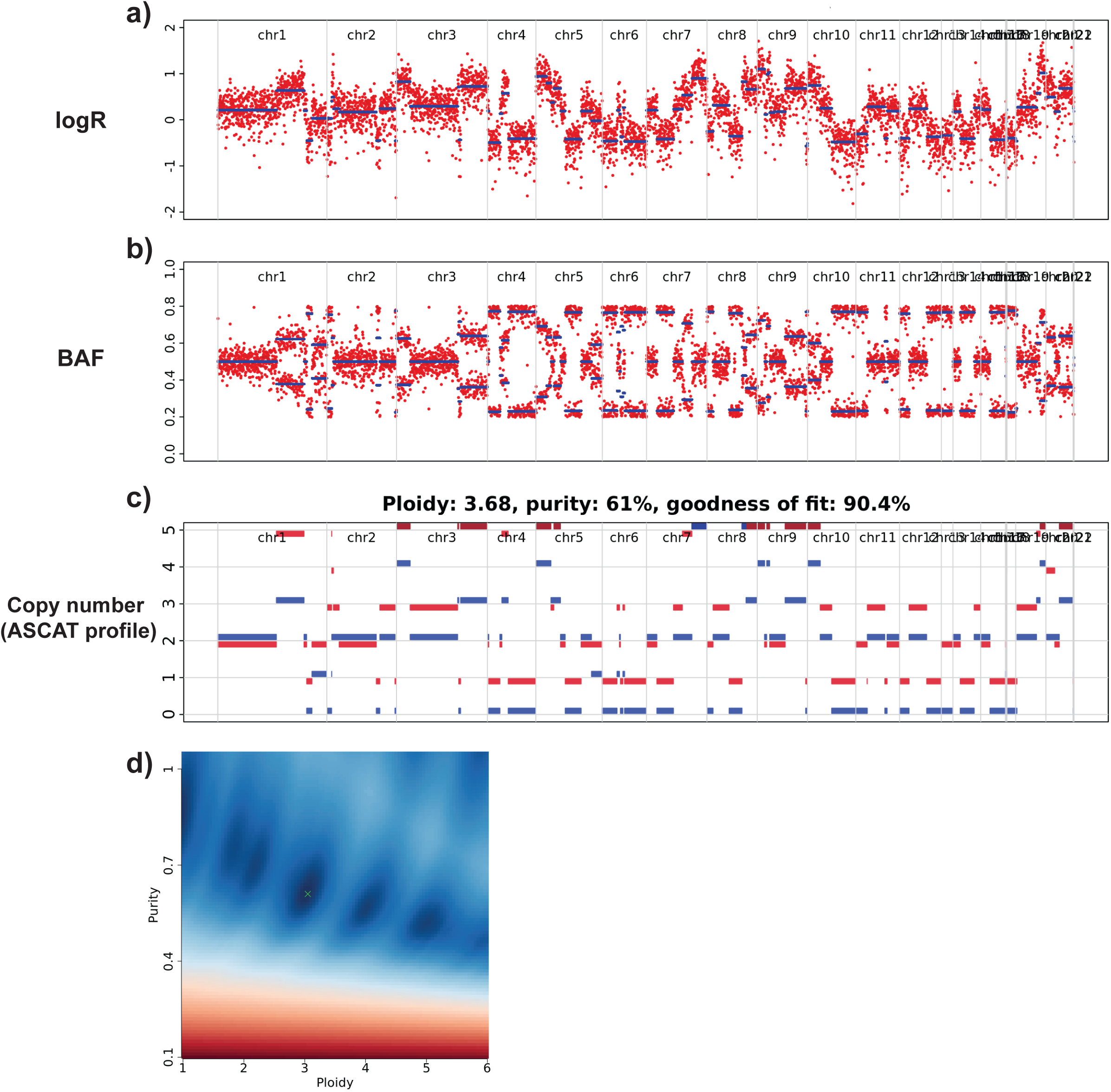
Plots generated by the HRD scoring estimation pipeline for a given sample. **a, b)** The LogR and BAF tracks provide cleaned genome-wide signals of a sample’s copy number variation. **c)** From these tracks, the allele-specific copy number profile is generated by ASCAT and the major and minor copy number across the genome are summarised in two different colours (blue and red). **d)** In addition, the estimation of the tumour purity and ploidy made by ASCAT is plotted in the sunrise plot.

## Survey for the fully-automated integrative workflow

Instructions:

Please rate the following statements based on your experience with the new automated integration workflow. Use the scale from 1 to 5, where:

1 = Strongly Disagree
2 = Disagree
3 = Neutral
4 = Agree
5 = Strongly Agree

### Workflow Impact & Results Accessibility

1. I appreciate that the pipeline runs automatically without requiring manual input or script execution.
2. The automated nature of the pipeline reduces my need to interact with tools or command-line interfaces.
3. I clearly understand when the results are generated and made available.

### Time & Workflow Efficiency

1. The automation of previously manual tasks is effective and reliable.
2. The workflow eliminates redundant or repetitive steps I previously had to do.
3. The pipeline improves the overall turnaround time of somatic variant analysis and interpretation.

### Integration & Impact on Work

1. I spend less time troubleshooting or resolving errors compared to before.
2. I don’t have to monitor anymore the status of the individual analysis steps.
3. The fully-automated workflow has positively impacted my productivity.

### Overall Satisfaction

1. I am satisfied with the benefits of the fully-automated workflow.
2. The workflow is an improvement over the previous manual approach.
3. I would recommend this pipeline to colleagues in similar roles.

